# Rental housing may contribute to racial and ethnic disparities in upper respiratory infections

**DOI:** 10.64898/2026.05.13.26351511

**Authors:** Darlene Bhavnani, Peter Dunphy, Matthew Wilkinson, Adam L. Haber, Elizabeth C. Matsui

## Abstract

**Objective:** Upper respiratory infections (URI) are the major trigger of asthma exacerbations in children with asthma and are more likely to be reported by Black and Mexican American children compared to White children in the US. We aimed to evaluate the extent to which obesity, nicotine exposure, household size, and socioeconomic status (SES) explained this excess URI risk among all children and among children with asthma.

**Study Design:** Data collected on children aged 6-17 years from the National Health and Nutritional Examination Survey (2007-2012) were analyzed using survey weights and a mediation approach. Household SES was analyzed as a cumulative score reflecting income poverty ratio, education, and rental housing. URI was defined as cough, cold, phlegm, runny nose, or other respiratory illness (excluding hay fever and allergies) in the past 7 days.

**Results:** Obesity and serum cotinine, a marker of nicotine exposure, explained little to none of the excess risk of URI while SES explained 36.4% (95% CI=34.1-38.6) in Black and 28.5% (95% CI=26.7-30.5) in Mexican American children. Living in rental housing and income poverty ratio<2, explained half (49.6%, 95% CI=46.9-52.3) and 20% (19.7%, 95% CI=18.9-20.5) of the excess URI risk among Black children, respectively. In Mexican American children, rental housing and low educational attainment each explained approximately 15-17% of the excess URI risk. Results were comparable among children with asthma.

**Conclusions:** Markers of poverty, such as rental housing, contributed substantially to the excess risk of URI among Black and Mexican American children, including among those with asthma.

## Introduction

School-aged children have on average 6-8 upper respiratory viral infections per year and in any given week, approximately one in five children have an upper respiratory infection (URI).^1–4^ URIs can lead to substantial loss of learning and income stemming from missed school and missed work.^2^ Using National Health and Nutrition Examination Survey (NHANES) data from across the US, we reported that Black and Mexican American children are 30-50% more likely to have URIs than White children.^4^ Among those with asthma, Black and Mexican American children are twice as likely to have URIs than White children. Although upper respiratory viral infections are the major trigger of asthma exacerbations in children with asthma,^5–8^ URIs have been overlooked as a potential contributor to racial and ethnic disparities in asthma exacerbations.^9^ It is unclear why Black and Mexican American children are at higher risk of acquiring a URI.

There are several possible explanations for this observed difference in the risk of URI: 1) differential exposure to upper respiratory viruses and or 2) differential efficacy of host-pathogen defense pathways that may cause an individual to be more susceptible to viral infections once exposed. There are a range of factors commonly found among historically marginalized populations (including Black and Mexican American populations) that may contribute to one or both mechanisms, some of which are captured in the NHANES dataset. Low socioeconomic status (SES) may relate to increased exposure to respiratory viruses through crowding and or poor ventilation indoors.^10–13^ Obesity, nicotine exposure, and the physiological toll of having low SES may impair host-pathogen defense mechanisms, potentially increasing an individual’s susceptibility to URI.^14–18^ We evaluated the extent to which these individual and household factors explained URI disparities between Black, Mexican American, and White children in the general population and among children with asthma.

## Methods

### Data collection and variables

Data were collected on children aged 6-17 years in NHANES. Data collected from 2007-2012 were selected for analysis because they were the most recent years to have included specific questions about upper respiratory illness. NHANES employs a complex, multistage, clustered design to collect data on a representative sample of the noninstitutionalized population in the US. Data were collected through interviews, examinations, and laboratory tests, and were released in 2-year survey cycles.^19^ Two-year examination sampling weights were divided by 3, representing the number of pooled survey cycles. Parental consent was obtained for all children and child assent was obtained from children aged 7-17 years.^20^ Data collection protocols were approved by the National Center for Health Statistics Research ethics review board.

Race and ethnicity were self-reported. Children were reported by NHANES as non-Hispanic White (henceforth, “White”), non-Hispanic Black (henceforth, “Black”), Mexican American, other Hispanic, or other race. We examined race and ethnicity in this analysis as a social construct. Current asthma was ascertained from the combination of two standardly used questions,^21^ “Has a doctor or other health professional ever told you that you have asthma?” and “Do you still have asthma?” URI was captured from a question about having a cough, cold, phlegm, runny nose, or other respiratory illness, excluding allergies or hay fever, in the past 7 days. Parents responded for children aged 6-11 years and children aged 12-17 years responded for themselves. Body mass index (BMI) was calculated from weight and height. Consistent with American Academy of Pediatrics recommendations^22^ and using CDC reference equations in the childsds package in R,^23^ children with a sex- and age-specific BMI in the 95^th^ percentile or higher were categorized as having obesity. Serum cotinine, a metabolite of nicotine, was used as a biomarker of nicotine exposure. Household data included household size, education, income poverty ratio, and home ownership. Education referred to the highest level of education completed by the household reference person (the first adult listed on the screening roster who owned or rented the residence).^19^ Education was recoded as 0 (completed more than high school), 1 (completed high school), or 2 (less than high school). Income poverty ratio, which refers to the ratio of family income to the Department of Health and Human Services poverty guidelines, was recoded as 0 (≥2) or 1 (<2). Home ownership was recoded as 0 (owned or being bought) or 1 (rental or some other arrangement). Given that other arrangements accounted for less than 2% of responses in the latter category, we refer to this category as rental. Education, income poverty ratio, and home ownership were examined separately and as a single integrated measure of SES. Although there is not a consistent integrated measure of SES in the literature, previous studies have used measures that capture multiple domains of SES, including education and income.^24,25^ In this study, SES was measured using a cumulative score from 0-4; higher scores corresponded to higher levels of disadvantage. Inclusion of health insurance in the SES score was considered; however, nearly all children in the US have health insurance^26^ and thus, this variable was not considered informative.

#### Data analysis

We included White, Black, and Mexican American children with complete observations. We used survey-weighted logistic regression models to estimate the association between potential mediators (obesity, current asthma, serum cotinine, SES score, and component variables of the SES score) and URI. All models were adjusted for age category (6-8, 9-11 and 12-17 years) and sex^4^ and fit using the survey package in R.^27^

To evaluate the proportion of the association between race/ethnicity and URI explained by each mediator, we estimated average causal mediation effects (i.e., the indirect effects of race/ethnicity via each mediator) using the mediation package in R.^28^ We fit survey weighted logistic regression models (adjusted for age and sex) for each mediator and URI, simulated model parameters from their sampling distribution, simulated potential values of mediators, and simulated values of URI (presence/absence), given the simulated values of mediators. The proportion of the association between race/ethnicity and URI explained by a mediator was estimated as the average causal mediation effect divided by the total effect of race/ethnicity on URI.^29^

## Results

In our sample of the general population, there were 3,510 children aged 6-17 years with complete observations. Accounting for survey weights, the population was 66.7% White, 16.3% Black, and 17.1% Mexican American. Approximately one in five children reported a URI in the past week (19.8%) and one in five children had obesity (21.3%, Table 1). Characteristics of children included in the study were similar to those excluded from the study (N=1,790, Supplemental Results, eTable 1). The prevalence of URI was 22.4% (95% CI = 19.2-26.0) in Black, 25.4% (95% CI = 22.5-28.6) in Mexican American, and 17.7% (95% CI = 14.9-21.0) in White children. In households with Black, Mexican American and White children, the median socioeconomic score was 2 (IQR = 1-3), 3 (IQR = 2-4), and 1 (IQR=0-2), respectively. Notably, while approximately a quarter (23.1%) of White children lived in rental housing, nearly half (46.3%) of Mexican American children and nearly two-thirds (63.1%) of Black children lived in rental housing.

**Table 1.** Characteristics of the study population (NHANES, 2007-2012)

|  | Survey weighted percent or median (IQR) |  |  |  |
| --- | --- | --- | --- | --- |
|  | All | White | Black | Mexican American |
| **Children in the general population (N=3,510)** |  |  |  |  |
| Age, years | 12 (9-15) | 12 (9-15) | 12 (9-15) | 11 (8-14) |
| Male | 50.6 | 50.6 | 50.1 | 50.9 |
| Reported a URI<sup>a</sup> | 19.8 | 17.7 | 22.4 | 25.4 |
| Obesity | 21.3 | 18.6 | 25.0 | 28.0 |
| Current asthma | 12.2 | 11.8 | 18.3 | 7.9 |
| Serum cotinine, ng/mL | 0.04<br>(0.01-0.26) | 0.04<br>(0.01-0.32) | 0.10<br>(0.03, 0.59) | 0.02<br>(0.01, 0.05) |
| Household Size | 4 (4-5) | 4 (4-5) | 4 (3-5) | 5 (4-6) |
| Socioeconomic score | 1 (0-2) | 1 (0-2) | 2 (1-3) | 3 (2-4) |
| Income <2x the poverty line | 46.1 | 34.2 | 64.9 | 74.8 |
| Education score | 0 (0-1) | 0 (0-1) | 0 (0-1) | 2 (0-2) |
| More than high school (0) | 57.5 | 67.0 | 51.8 | 25.6 |
| High school (1) | 21.6 | 21.0 | 25.2 | 20.4 |
| Less than high school (2) | 20.9 | 11.9 | 23.0 | 54.0 |
| Lives in rented housing | 33.6 | 23.1 | 63.1 | 46.3 |
| **Children with current asthma (N=433)** |  |  |  |  |
| Age, years | 13 (10-15) | 13 (10-15) | 12 (8-14) | 11 (8-14) |
| Male | 53.7 | 50.4 | 66.1 | 45.6 |
| Reported of a URI<sup>a</sup> | 22.5 | 18.1 | 29.8 | 32.2 |
| Obesity | 27.5 | 25.2 | 31.9 | 31.0 |
| Serum cotinine, ng/mL | 0.06<br>(0.02-0.64) | 0.07<br>(0.01-0.82) | 0.13<br>(0.03-0.70) | 0.02<br>(0.01-0.08) |
| Household Size | 4 (4-5) | 4 (4-5) | 4 (3-5) | 5 (4-6) |
| Socioeconomic score | 1 (0-3) | 1 (0-2) | 2 (1-3) | 2 (1-3) |
| Income <2x the federal poverty level | 50.4 | 41.1 | 69.5 | 62.5 |
| Education score | 0 (0-1) | 0 (0-1) | 0 (0-2) | 1 (0-2) |
| More than high school (0) | 54.8 | 57.9 | 50.4 | 46.4 |
| High school (1) | 23.3 | 23.7 | 22.6 | 22.1 |
| Less than high school (2) | 21.9 | 18.4 | 27.0 | 31.5 |
| Lives in rental housing | 39.0 | 29.7 | 65.0 | 36.0 |

<sup>a</sup>Upper respiratory infection

There were 433 children with current asthma in the sample, equaling one in eight children (12.2%). Prevalence of current asthma was 18.3% in Black children, 7.9% in Mexican American children and 11.8% in White children. Among children with asthma, 22.5% had a URI and 27.5% had obesity. Racial and ethnic differences in URI and SES scores in the general population sample were also observed in children with asthma. Participants with asthma and excluded from the analysis due to missing data were younger (median age was 11 years compared to 13 years), had a higher prevalence of URI (27.3%), obesity (36.6%), and had a higher SES score (2 vs.1) than those with asthma and included in the analysis.

In the general population, we observed positive associations between obesity and SES score and URI (Table 2). Children with obesity had a 43% greater odds of reporting a URI compared to children without obesity (OR=1.43, 95% CI=1.14-1.81). As SES score increased by a unit of one (suggesting lower access to financial and educational resources), the odds of a URI increased by 15% (OR=1.15, 95% CI=1.06-1.24). Component variables of the SES score were also significantly associated with a URI. For example, living in rental housing was associated with a 53% increase in the odds of a URI (OR=1.53, 95% CI = 1.18-1.97).

**Table 2.** Associations between potential mediators and upper respiratory infection in the general population of children (N=3,510)

| Mediator | Survey weighted percent or median (95% CI) |  | Survey weighted logistic regression<sup>a</sup> |
| --- | --- | --- | --- |
|  | No URI | URI | OR (95% CI) |
| Obesity | 20.1 (18.3, 22.2) | 25.9 (22.2, 29.8) | 1.43 (1.14, 1.81) |
| Current asthma | 11.8 (10.3, 13.5) | 13.9 (10.9, 17.5) | 1.15 (0.84, 1.56) |
| Serum Cotinine, ng/mL | 0.04 (0.03, 0.05) | 0.05 (0.03, 0.06) | 1.04 (0.997, 1.08) |
| Household size | 4 (4, 5) | 4 (4, 5) | 0.98 (0.89, 1.07) |
| SES score | 1 (1, 2) | 1 (1,2) | 1.15 (1.06, 1.24) |
| Income <2x the federal poverty level | 45.2 (41.2, 49.3) | 49.6 (45.1, 54.1) | 1.27 (1.05, 1.52) |
| Education score | 0 (0, 1) | 0 (0, 1) | 1.16 (1.01, 1.34) |
| Living in rental housing | 31.8 (27.5, 36.5) | 40.5 (35.0, 46.2) | 1.53 (1.18, 1.97) |

<sup>a</sup>Each model was adjusted for age category and sex.

Among children with asthma, SES score and two components of the score (income poverty ratio <2 and living in rental housing) were significantly associated with URI (Table 3). For example, living in rental housing was associated with approximately 160% increase in the odds of a URI among children with asthma (OR=2.57, 95% CI=1.38-4.80).

**Table 3.** Associations between potential mediators and upper respiratory infection in the children with current asthma (N=433).

| Mediator | Survey weighted percent or weighted median (95 % CI) |  | Survey weighted logistic regression<sup>a</sup> |
| --- | --- | --- | --- |
|  | No URI | URI | OR (95% CI) |
| Obese | 25.2 (19.1, 32.4) | 35.5 (24.9, 47.7) | 1.67 (0.93, 3.01) |
| Serum Cotinine, ng/mL | 0.05 (0.03, 0.14) | 0.09 (0.05, 0.32) | 1.07 (0.99, 1.16) |
| Household size | 4 (4, 5) | 4 (4, 5) | 1.03 (0.81, 1.32) |
| SES score | 1 (1, 2) | 2 (2, 3) | 1.31 (1.04, 1.64) |
| Income <2x the federal poverty level | 46.7 (35.9, 57.9) | 62.9 (45.5, 77.6) | 2.11 (1.09, 4.08) |
| Education score | 0 (0, 1) | 0 (0, 1) | 1.08 (0.75, 1.55) |
| Living in rental housing | 34.1(25.1, 44.5) | 55.6 (41.0, 69.4) | 2.57 (1.38, 4.80) |

<sup>a</sup>Each model was adjusted for age category and sex.

In the general population of children, SES score explained 29-36% of the association between Black and Mexican American identity and URI (Figure 1 and Supplemental Results, eTable 2). In a separate model, living in rental housing, explained half of the association between Black identity and URI (49.6%, 95% CI=46.9-52.3). Living in rental housing explained a lower proportion of the association between Mexican American identity and URI (17.4%, 95% CI=16.6-18.3). Serum cotinine concentrations explained ∼10% of the association between Black identity and URI. In contrast, negative mediation by serum cotinine was observed for Mexican American children, suggesting that the lower concentrations of serum cotinine observed across the population of Mexican American children are associated with a lower risk of URI relative to White children. Negative mediation was also observed by household size for Mexican American children. This suggests that larger household sizes observed among Mexican American compared to White families contribute to a lower risk of URI relative to White children. Obesity and asthma explained little to none of the association between Black or Mexican American identity and URI.

**Figure 1.**
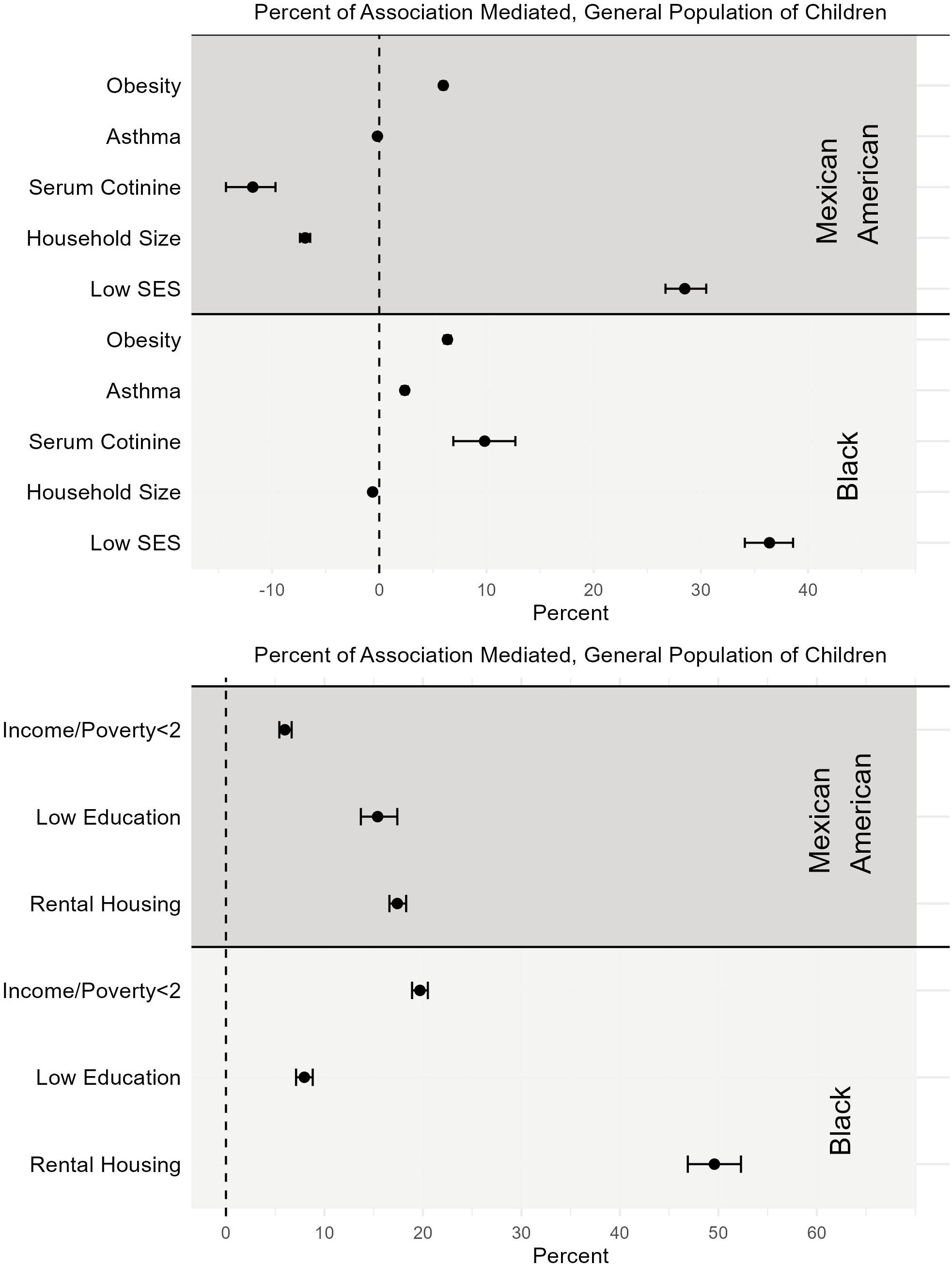
Percent of the association between racial and ethnic identity (reference = White) and upper respiratory infection explained by potential mediators (top) and specific components of the socioeconomic score (bottom) in the general population of children.

Among children with asthma, predominant mediators of the association between Black identity and URI were SES score (21.5%, 95% CI= 16.0-28.7), income poverty ratio <2 (27.8%, 95% CI=21.1-36.6) and living in rental housing (37.5%, 95% CI=25.2-55.3, Figure 2 and Supplemental Results, eTable 3). Among Mexican American children, important mediators were SES score (18.1%, 95% CI=11.7-26.7) and income poverty ratio <2 (17.1, 95% CI=11.2-26.2). Serum cotinine results were qualitatively similar to those in the general population however, wide confidence intervals owing to a large variability in concentrations among White and Black children with asthma may have precluded our ability to detect mediation with statistical significance.

**Figure 2.**
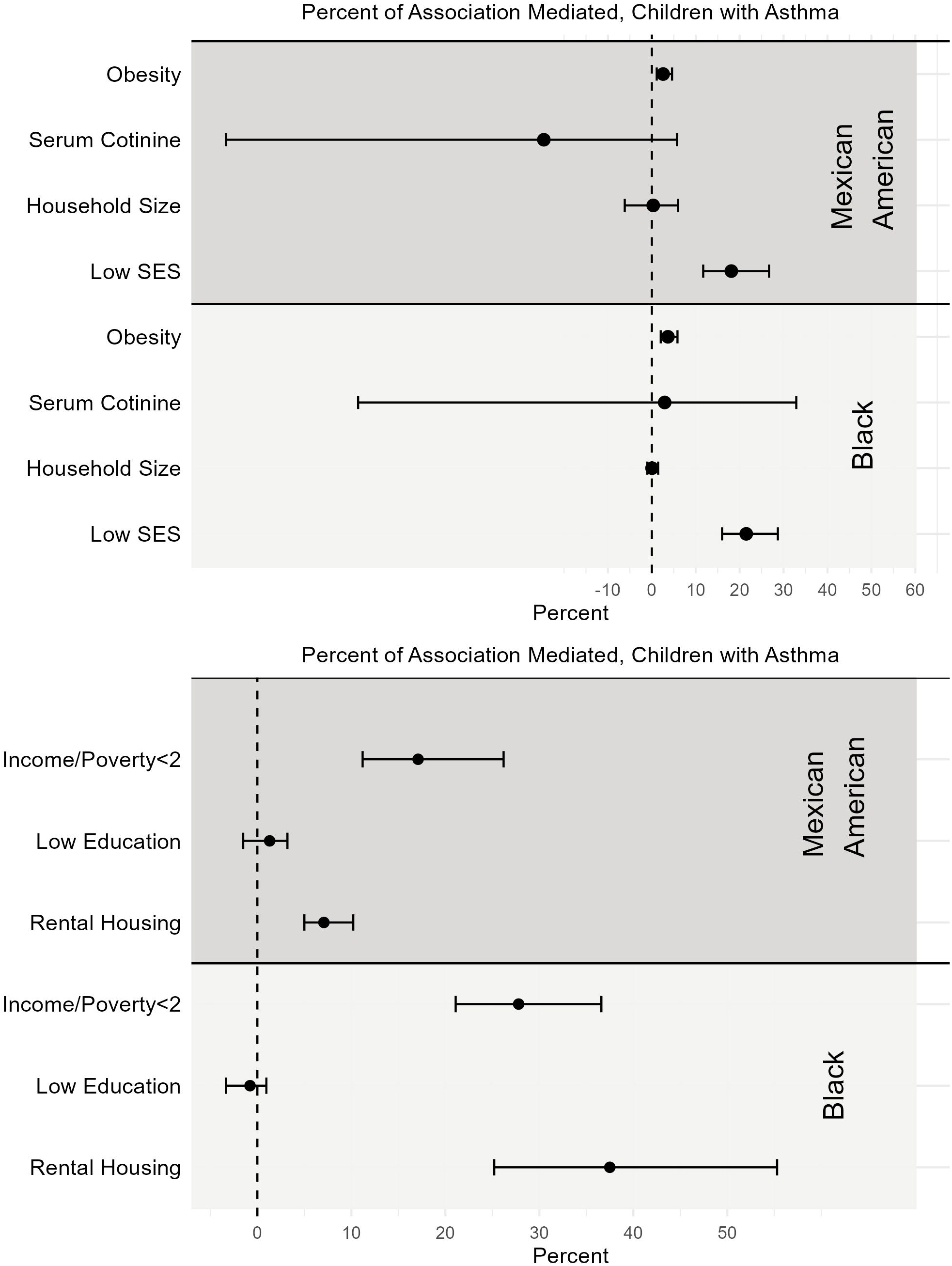
Percent of the association between racial and ethnic identity (reference = White) and upper respiratory infection explained by potential mediators (top) and specific components of the socioeconomic score (bottom) in children with asthma.

## Discussion

In this nationally representative sample, SES score and its component factors explained a substantial portion of the disparity in URI between Black and Mexican American children and White children. Having lower SES explained 29-36% of URI disparities in the general population of children and 18-22% of URI disparities in children with asthma. These findings suggest that low SES or poverty, more prevalent among Black and Mexican Americans, may contribute to the excess burden of URI. URIs in children can lead to 189 million missed school days and 126 million missed workdays among caregivers each year.^2^ Not only does a disproportionate share of this social and economic burden of URIs fall on Black and Mexican American families, but these families already have lower incomes, limited employment benefits, less sick leave, and fewer educational opportunities,^30,31^ potentially amplifying poverty levels and leading to a circular relationship between poverty and URI.

The implications of these disparities in URI may be greater in children with asthma, as upper respiratory viruses trigger up to 85% of asthma exacerbations.^5–8^ While factors including health care access, differential use of controller medications, air pollution, allergens and other characteristics of the physical environment have been identified as causes of racial and ethnic disparities in asthma morbidity,^32,33^ the contribution of upper respiratory viral infections to these disparities has been underappreciated. Understanding that contextual factors, like SES, play a large role in disparities in upper respiratory infection between Black, Mexican American, and White children with asthma may inform approaches to address racial and ethnic differences in asthma exacerbations.

Living in rental housing was associated with the risk of a URI and explained a significant proportion of the excess risk of URI among Black and Mexican children. The association between living in rental housing and URI is consistent with findings from a series of carefully designed viral challenge studies in adults. In one of these studies, Cohen et al^17^ found that risk of infection with a common cold virus (and separately, risk of a symptomatic infection) increased as the number of childhood years spent in an owned home decreased. Cohen et al^18^ later observed that shorter CD8+ CD28-T-cell telomere length (indicating that the cells are approaching senescence) helped to explain these associations. Immune pathways associated with 1) initial host viral defense and 2) the local inflammatory response giving rise to cold symptoms, may be involved.^18^ More recently, Cohen et al.^34^ reported that positive relationships with parents during childhood buffered the effect of home ownership on symptomatic infections. The results of our study in the context of those previously conducted by Cohen’s group suggest that a lack of home ownership during childhood may lead to an impairment of the immune response to upper respiratory viruses in childhood as well, and this impairment may be modifiable by social factors.

Our observation that markers of SES, like a lack of home ownership, explained disparities in the risk of URI among children with asthma means that they may also explain disparities in the risk of viral-associated asthma exacerbations. In one study a lack of home ownership combined with greater material hardship helped to explain the disparity in asthma exacerbations between Black and non-Hispanic White children with asthma.^35^ However, this study did not differentiate between viral-associated and non-viral asthma exacerbations. Furthermore, the causal mechanisms underlying the relationship between living in rental housing and viral-associated asthma exacerbations require further examination. These mechanisms may not include crowding in the home, given our observation that household size was not associated with URI, and could include psychological stress associated with financial hardship. Psychological stress is strongly associated with an increased risk of URI.^36^

Alternatively, the mechanisms could include environmental factors that children in rental housing may be more exposed to. Children living in public housing are exposed to higher concentrations of air pollution,^37^ which is linked to impaired host viral defense mechanisms.^16^ Similarly, children living in rental homes may have insufficient heating, potentially leading to an increased exposure to cold temperatures and impaired viral defense.^38–40^ Indoor pest allergens are more commonly found in rental housing,^41^ and among children with asthma, exposure to higher concentrations of indoor pest allergens may increase the risk of symptomatic upper respiratory viral infections.^42^ These environmental factors are also associated with non-viral asthma exacerbations,^43,44^ which could mean that there is an alternate pathway through which disparities in asthma exacerbations result from living in rental housing, potentially leading to a double hit on home renters.

While the contribution of low SES to URI disparities may be large, other factors in the home, school, and neighborhood environment may be important. We found that nicotine exposure helped to explain the excess risk of URI among Black children. Additionally, low SES may be a proxy for other exposures commonly found in neighborhoods with a high proportion of renters. For example, there may be increased exposure to URI in schools, owing to lack of sick leave among parents in the neighborhood, poor ventilation in classrooms, and or crowding.^13^ Other factors could include neighborhood-level disadvantage, which in some cases, is measured using the proportion of owned homes in a neighborhood,^45^ and is associated with viral infection^46–48^ and racial and ethnic disparities in asthma.^33^ At larger spatial scales, outdoor air pollution is correlated with viral risk^49,50^ and at the neighborhood level, outdoor air pollution may help to explain racial and ethnic disparities in asthma.^33,51^ Given the conceptual link between living in rental housing and these neighborhood-level factors associated with risk of respiratory disease, studies that can help to tease out the effects of these highly correlated factors could provide insight into specific targets for intervention.

There are several limitations of this study. As previously discussed,^4^ our reliance on self-report of URI as a marker of upper respiratory viral infection may have led to outcome misclassification. Nevertheless, prior studies have detected upper respiratory viruses in 60-66% of children reporting a URI,^1,52^ lending confidence to the use of survey data. Additionally, report of a URI is expected to reflect viral infection similarly by race, ethnicity, and socioeconomic status,^1,52^ suggesting that outcome misclassification is likely to be non-differential and more likely to bias results towards the null. NHANES included this question about URI from 2007 to 2012. While URI data have not been collected since, more recent reports of racial/ethnic disparities in respiratory viral infection^53^ and associations between poverty and URI,^48^ support the current relevance of our findings. Additionally, there is the potential for selection bias stemming from missing data and the smaller sample size of children with asthma that may have limited our ability to detect statistically significant associations. However, similar findings in the general population of children, which was not constrained by sample size, help to bolster our conclusions about the population of children with asthma. Another limitation is that the mediation approach relies on correct specification of the regression models. Although we accounted for confounding by age category and sex, there may be unmeasured confounders that were not accounted for, such as seasonality which was not available in our dataset, that may have resulted in model misspecification. A major strength of this study is the use of nationally representative data collected by oversampling Black and Mexican American populations. This allowed us to examine mediation separately among groups, which was critical to learning that there are different factors that explain disparities in URI between Black, Mexican American and White children.

Low SES appears to be a major contributor to the excess risk of URI among Black and Mexican American children, including among those with asthma. Collectively, these observations implicate aspects of poverty in the major public health burden of URIs in Black and Mexican American populations – accounting for approximately one third of the excess burden of URIs in Black and Mexican American children and approximately one fifth of the excess burden of viral-associated asthma exacerbations in Black and Mexican American children with asthma. The substantial contribution made by one specific indicator of poverty - living in rental housing - to the excess burden of URIs in both Black and Mexican American children may lend insight into the underlying mechanisms through which low SES leads to disparities in URI. It may also inform future studies of the mechanisms behind the association between low SES and disparities in URI, which could help to identify targets for interventions to reduce these disparities.

## Data Availability

These data are made publicly available by the Centers for Disease Control and Prevention National Center for Health Statistics.

## Acknowledgements

We would like to thank Dr. Paul J. Rathouz for his statistical advice and mentorship.

## Notes

**Funding**: Research was supported by the National Center for Advancing Translational Sciences, National Institutes of Health, through Grant KL2 TR002646. Effort on this project was also supported by core funds of the Dell Medical School at the University of Texas at Austin (Bhavnani) and the National Institutes of Health (grants R01ES035131[Matsui], R01ES023447 [Matsui] and R01ES034803 [Matsui]). The content is solely the responsibility of the authors and does not necessarily represent the official views of the NIH.

The authors declare no conflicts of interest. AI tools were not used in the development of this manuscript.

### Competing Interest Statement

The authors have declared no competing interest.

### Funding Statement

Research was supported by the National Center for Advancing Translational Sciences, National Institutes of Health, through Grant KL2 TR002646. Effort on this project was also supported by core funds of the Dell Medical School at the University of Texas at Austin (Bhavnani) and the National Institutes of Health (grants R01ES035131[Matsui], R01ES023447 [Matsui] and R01ES034803 [Matsui]). The content is solely the responsibility of the authors and does not necessarily represent the official views of the NIH.

