## Supplemental for "Rental housing may contribute to racial and ethnic disparities in upper respiratory infections"

**Supplemental Results**

**eTable 1**. Characteristics of children with and without missing data (NHANES, 2007-2012)

|  | Survey weighted percent or weighted median (IQR) | |
| --- | --- | --- |
| Children in the general population | Participants without missing data (N=3,510) | Participants with missing data (N=1,790) |
| Racial and ethnic identity |  |  |
| White | 66.7 | 66.6 |
| Black | 16.3 | 18.3 |
| Mexican American | 17.1 | 15.2 |
| Age, years | 12 (9-15) | 11 (8-14) |
| Male | 50.6 | 50.2 |
| Reported a URI^a^ | 19.8 | 19.9 |
| Obesity | 21.3 | 21.3 |
| Current asthma | 12.2 | 12.0 |
| Serum cotinine, ng/mL | 0.04 (0.01-0.26) | 0.04 (0.02-0.37) |
| Household Size | 4 (4-5) | 4 (4-5) |
| Socioeconomic score | 1 (0-2) | 1 (0-2) |
| Income less than 2x the poverty level | 46.1 | 42.5 |
| Education score | 0 (0-1) | 0 (0-1) |
| More than high school (0) | 57.5 | 55.7 |
| High school (1) | 21.6 | 23.9 |
| Less than high school (2) | 20.9 | 20.4 |
| Living in rental housing | 33.6 | 31.2 |
| Children with current asthma | Participants without missing data (N=433) | Participants with missing data (N=185) |
| Age, years | 13 (10-15) | 11 (8-14) |
| Male | 53.7 | 50.2 |
| Reported of a URI^a^ | 22.5 | 27.3 |
| Obesity | 27.5 | 36.6 |
| Serum cotinine, ng/mL | 0.06 (0.02-0.64) | 0.09 (0.02-0.35) |
| Household Size | 4 (4-5) | 4 (4-5) |
| Socioeconomic score | 1 (0-3) | 2 (1-3) |
| Income less than 2x the poverty level | 50.4 | 53.4 |
| Education score | 0 (0-1) | 0 (0-1) |
| More than high school (0) | 54.8 | 43.0 |
| High school (1) | 23.3 | 35.3 |
| Less than high school (2) | 21.9 | 21.7 |
| Living in rental housing | 39.0 | 36.6 |

^a^Upper respiratory infection

**eTable 2**. Results of the mediation analysis in the general population of children (N=3,510)

|  | Percent of the association explained (95% CI) | |
| --- | --- | --- |
| Mediator | Black vs. White | Mexican American vs. White |
| Obesity | 6.36 (6.06,6.67) | 5.97 (5.72, 6.22) |
| Asthma | 2.38 (2.11, 2.65) | -0.17 (-0.26, -0.07) |
| Serum Cotinine, ng/mL | 9.83 (6.91, 12.7) | -11.8 (-14.3, -9.68) |
| Household Size | -0.62 (-0.77, 0.47) | -6.91 (-7.40, -6.44) |
| Socioeconomic score | 36.4 (34.1, 38.6) | 28.5 (26.7, 30.5) |
| Income poverty ratio <2 | 19.7 (18.9, 20.5) | 5.98 (5.42, 6.67) |
| Education | 7.96 (7.12, 8.81) | 15.4 (13.7, 17.4) |
| Living in rental housing | 49.6 (46.9, 52.3) | 17.4 (16.6, 18.3) |

**eTable 3**. Results of the mediation analysis in children with asthma (N=433)

|  | Percent of the association explained (95% CI) | |
| --- | --- | --- |
| Mediator | Black vs. White | Mexican American vs. White |
| Obesity | 3.66 (2.03, 5.84) | 2.57 (1.11, 4.60) |
| Serum Cotinine, ng/mL | 2.92 (-66.9, 32.9) | -24.6 (-97.0, 5.73) |
| Household Size | 0.05 (-1.04, 1.44) | 0.31 (-6.17, 5.96) |
| Socioeconomic score | 21.5 (16.0, 28.7) | 18.1 (11.7, 26.7) |
| Income poverty ratio <2 | 27.8 (21.1, 36.6) | 17.1 (11.2, 26.2) |
| Education | -0.78 (-3.34, 0.96) | 1.31 (-1.51, 3.20) |
| Living in rental housing | 37.5 (25.2, 55.3) | 7.08 (5.00, 10.2) |
